# Significantly elevated antibody levels and neutralization titers in nursing home residents after SARS-CoV-2 BNT162b2 mRNA booster vaccination

**DOI:** 10.1101/2021.12.07.21267179

**Authors:** David H. Canaday, Oladayo A. Oyebanji, Elizabeth White, Debbie Keresztesy, Michael Payne, Dennis Wilk, Lenore Carias, Htin Aung, Kerri St. Denis, Maegan L. Sheehan, Sarah D. Berry, Cheryl M. Cameron, Mark J. Cameron, Brigid M. Wilson, Alejandro B. Balazs, Christopher L. King, Stefan Gravenstein

## Abstract

Nursing home (NH) residents have experienced significant morbidity and mortality to SARS-CoV-2 throughout the pandemic. Vaccines initially curbed NH resident morbidity and mortality, but antibody levels and protection have declined with time since vaccination, prompting introduction of booster vaccination. This study assesses humoral immune response to booster vaccination in 85 NH residents and 44 health care workers (HCW) that we have followed longitudinally since initial SARS-CoV-2 BNT162b2 mRNA vaccination. The findings reveal that booster vaccination significantly increased anti-spike, anti-receptor binding domain, and neutralization titers above the pre-booster levels in almost all NH residents and HCW to significantly higher levels than shortly after the completion of the initial vaccine series. These data support the CDC recommendation to offer vaccine boosters to HCWs and NH residents on an immunological basis. Notably, even the older, more frail and more multi-morbid NH residents have sizable antibody increases with boosting.

## Introduction

Nursing home (NH) residents completing the two-dose series of BNT162b2 mRNA vaccine dropped their antibody levels and neutralization titers by more than 80% in the 6 months following vaccination, and 57% no longer had detectable neutralization antibodies ^1^. Similarly, healthcare workers (HCWs) also experienced over an 80% decline, even if they had prior infection. This marked decline in antibody protection contributes to the increasing incidence of breakthrough SARS-CoV-2 infection in vaccinated individuals, especially among NH residents, that prompted the CDC’s recommendation and authorization of booster doses. Current reports on post-booster vaccination titers are limited to healthier older adults, excluding the more frail and multi-morbid NH population ^2,3^. In this study, we examined the effect of boosting on humoral immunity among NH residents and HCWs.

## Methods

Subjects who had previously provided serum samples to initial SARS-CoV-2 BNT162b2 mRNA vaccination series and also before and after booster were eligible for inclusion ^1,4^. Serum samples were obtained within 14 days before and 14±3 days after BNT162b2 mRNA booster. Study approval was obtained from the WCG Institutional Review Board. All subjects or their legally authorized representatives provided informed consent.

Immune response to the vaccine was assessed for IgG to spike protein (BAU/ml based on the Frederick National Laboratory standard which was calibrated to the WHO 20/136 standard) and its receptor binding domain (RBD) by bead-multiplex immunoassay using Wuhan strain ^1^. Stabilized full-length S protein (aa 16-1230, with furin site mutated) and RBD (aa 319-541) were conjugated to magnetic microbeads (Luminex) and Magpix assay system (BioRad, Inc). The mean fluorescent index was recorded after detecting antigen-specific IgG in patient serum using PE-conjugated Donkey F(ab)2 anti-human IgG, with Fcγ (Jackson Immunological).

To determine the neutralizing activity of vaccine recipients’ sera against coronaviruses, we produced lentiviral particles pseudotyped with spike protein based on the Wuhan strain as previously described ^5^. Briefly, neutralization assays were performed using a Fluent 780 liquid handler (Tecan) in 384-well plates (Grenier). Three-fold serial dilutions ranging from 1:12 to 1:8,748 were performed and added to 50–250 infectious units of pseudovirus for 1 hour. pNT50 values were calculated by taking the inverse of the 50% inhibitory concentration value for all samples with a pseudovirus neutralization value of 80% or higher at the highest concentration of serum. The lower limit of detection (LLD) of this assay is 1:12 dilution.

We assessed the geometric mean fold rise (GMFR) from pre- to post-boost and from 2 weeks post-initial vaccination to post-boost using a two-sided t-test on the log-transformed titer fold change values within each group. All p-values are presented without adjustment. All analyses were performed in R version 4.0.3.

## Results and Discussion

We sampled 85 NH residents (median age 77) and 44 HCWs (median age 50) (Table 1) from three NHs. We previously reported on this group for the 6-month period following completion of the initial SARS-CoV-2 BNT162b2 mRNA vaccination series ^1,4^. Booster vaccination significantly increased anti-spike, anti-RBD, and neutralization titers above the pre-booster levels in NH residents and HCWs, both in those with (“prior”) and without (“infection naive”) SARS-CoV-2 infection (p<0.001 in all groups) (Table 2, Fig. 1,2,3). Prior-infected NH residents, and infection-naive NH residents and HCWs all achieved higher anti-spike antibody and neutralization titers than observed 2 weeks after their initial vaccine series, demonstrating that boosting produced their highest lifetime titers to date (Table 2, p<0.02 in these groups). Infection-naive NH residents had the lowest initial vaccine response and lowest absolute titers 6 months later, but after boosting had a robust geometric mean fold rise (GMFR) of 9.3 in anti-spike antibody levels and a GMFR of 6 in neutralization titer from their prior peak titers 2 weeks after the initial vaccine series.

**Table 1.**
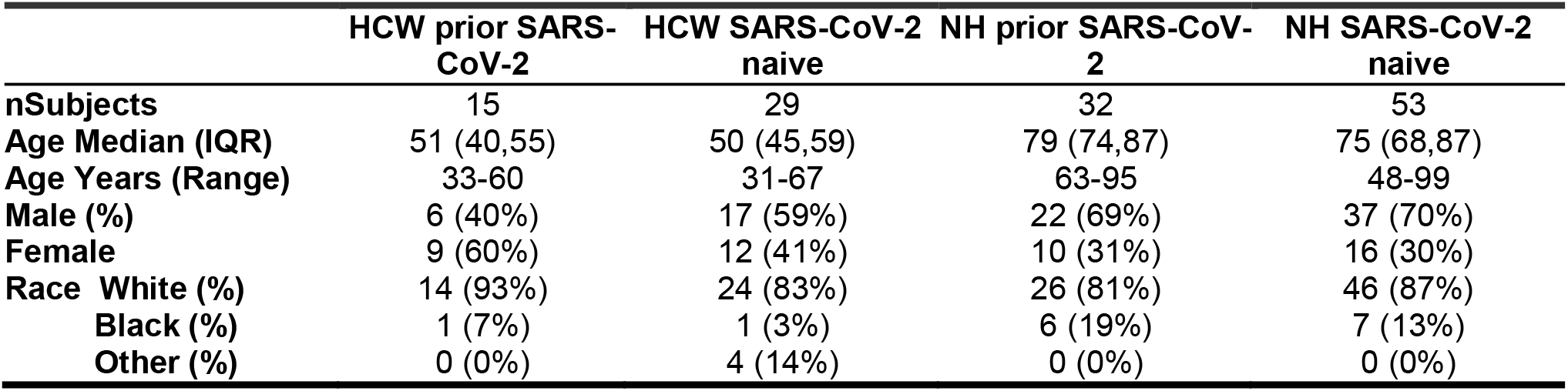
Subject Demographics.

**Table 2.**
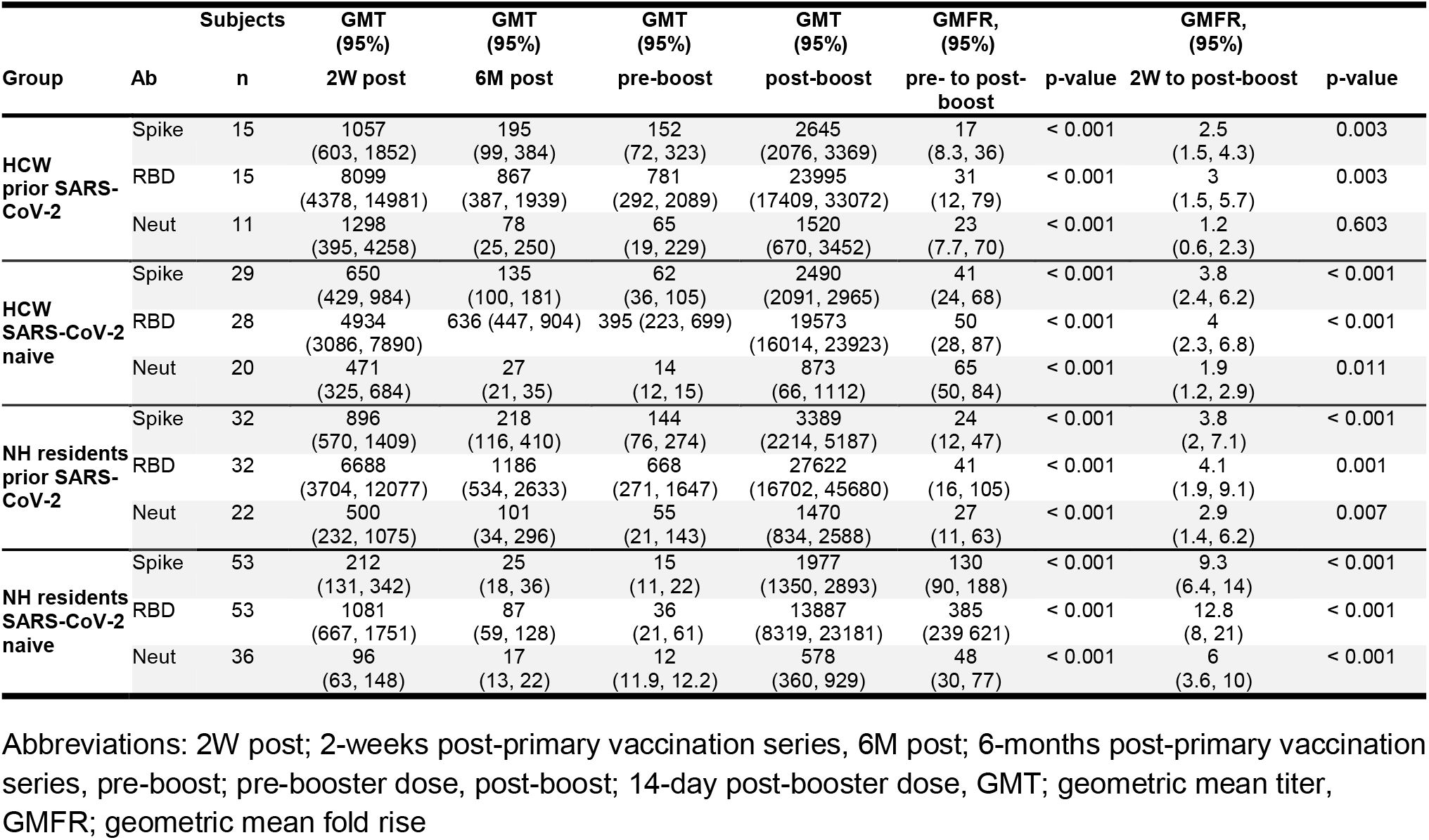
Antibody and neutralization titers.

**Figure 1.**
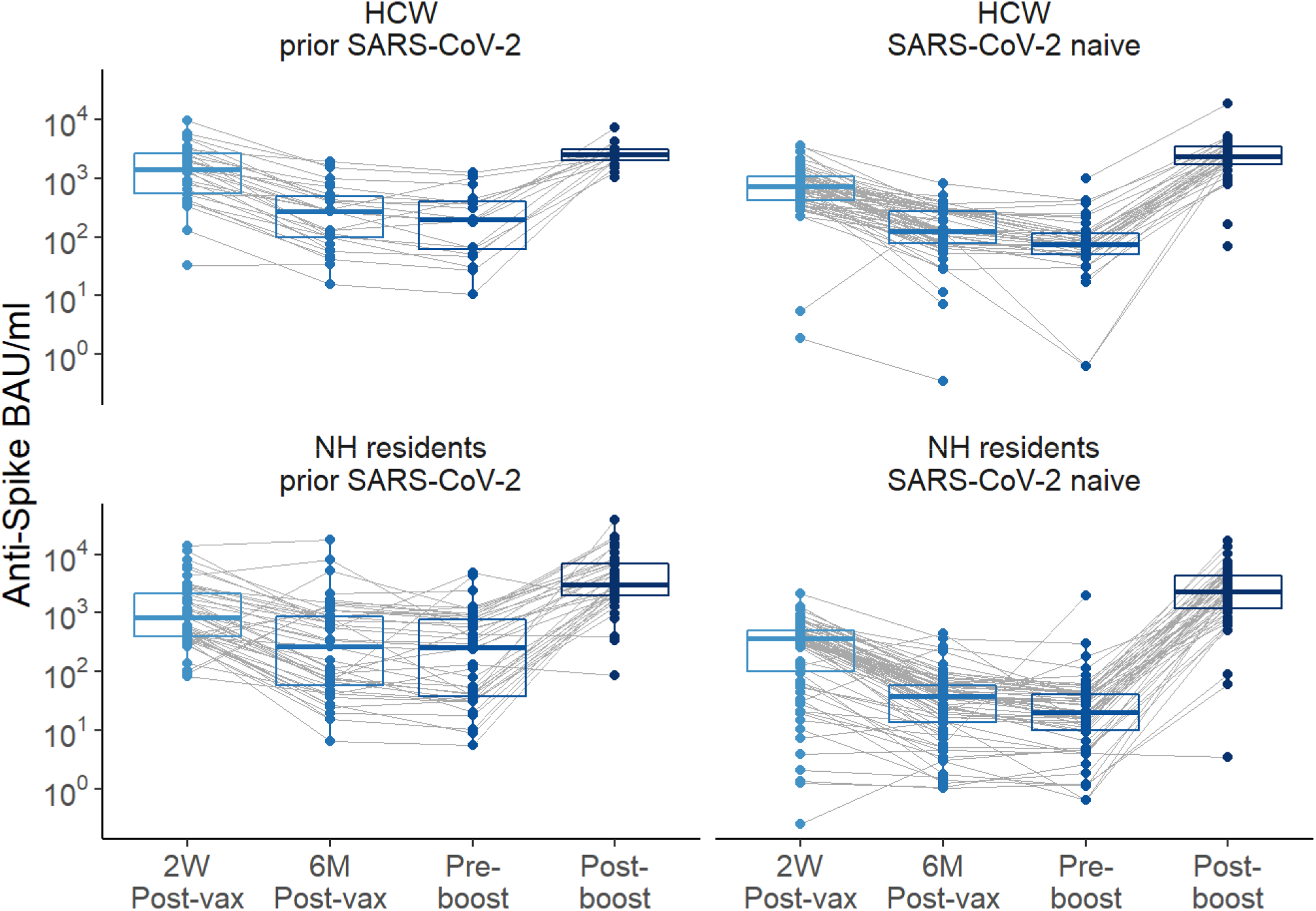
Anti-spike levels over time pre- and post-boost with BNT162b2 mRNA vaccination in healthcare workers (HCWs) and nursing home (NH) residents, with and without history of SARS-CoV-2. Anti-spike values depicted in the binding arbitrary units /milliliter (BAU/ml) based on the WHO standard. 2 weeks (2W Post-vax) and 6 months (6M Post-vax) post primary vaccination series and Pre-boost (generally 6-8 months after the first two-dose vaccination series) and Post-boost which is 14±3 days after vaccine boost.

**Figure 2.**
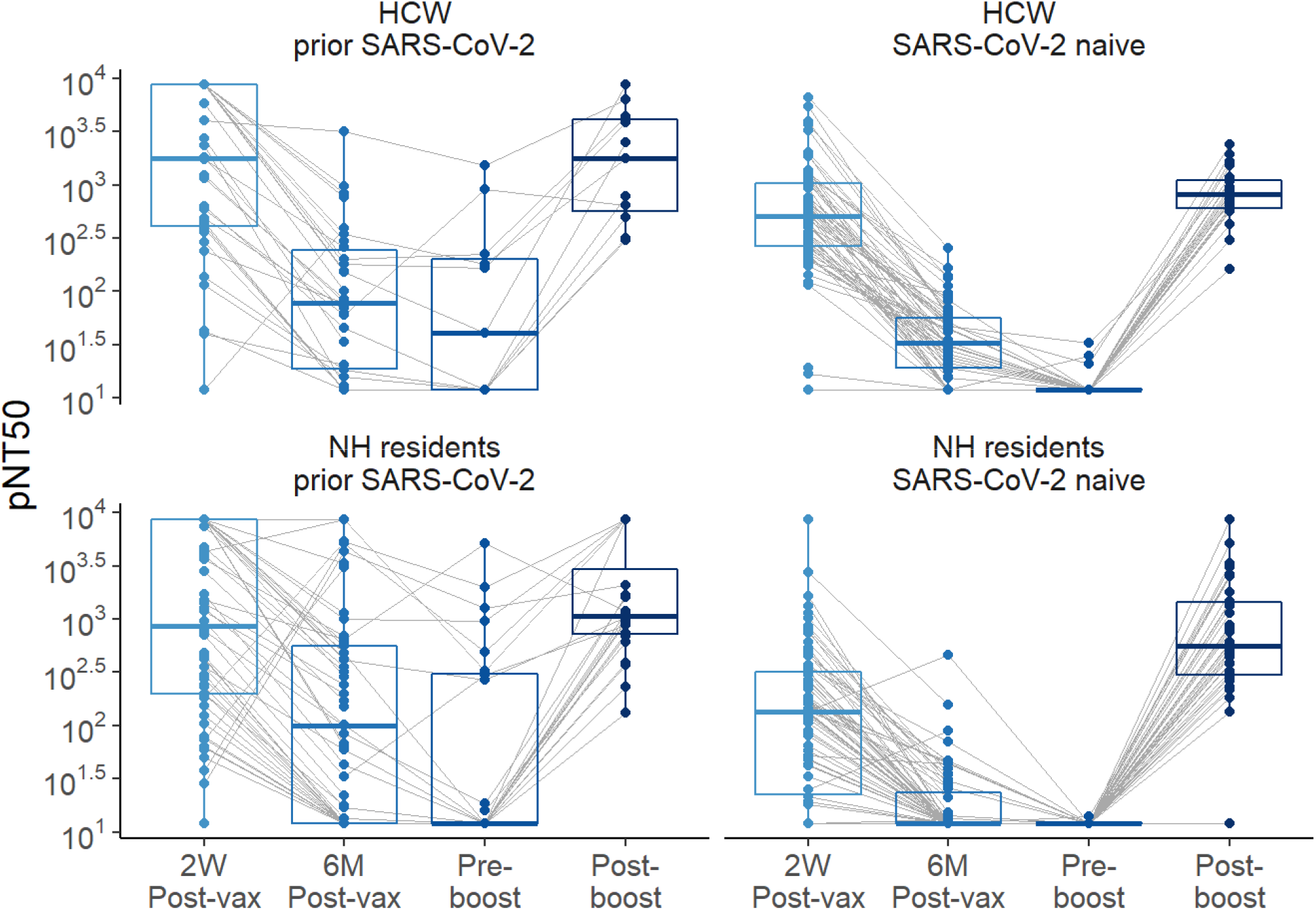
Neutralization titers over time pre- and post-boost with BNT162b2 mRNA vaccination in HCW and NH residents, with and without history of SARS-CoV-2. Pseudovirus neutralization (pNT50) values are shown. The upper limit of detection of the assay is 1:8748 and the low limit of detection of the neutralization assay is 1:12.

**Figure 3.**
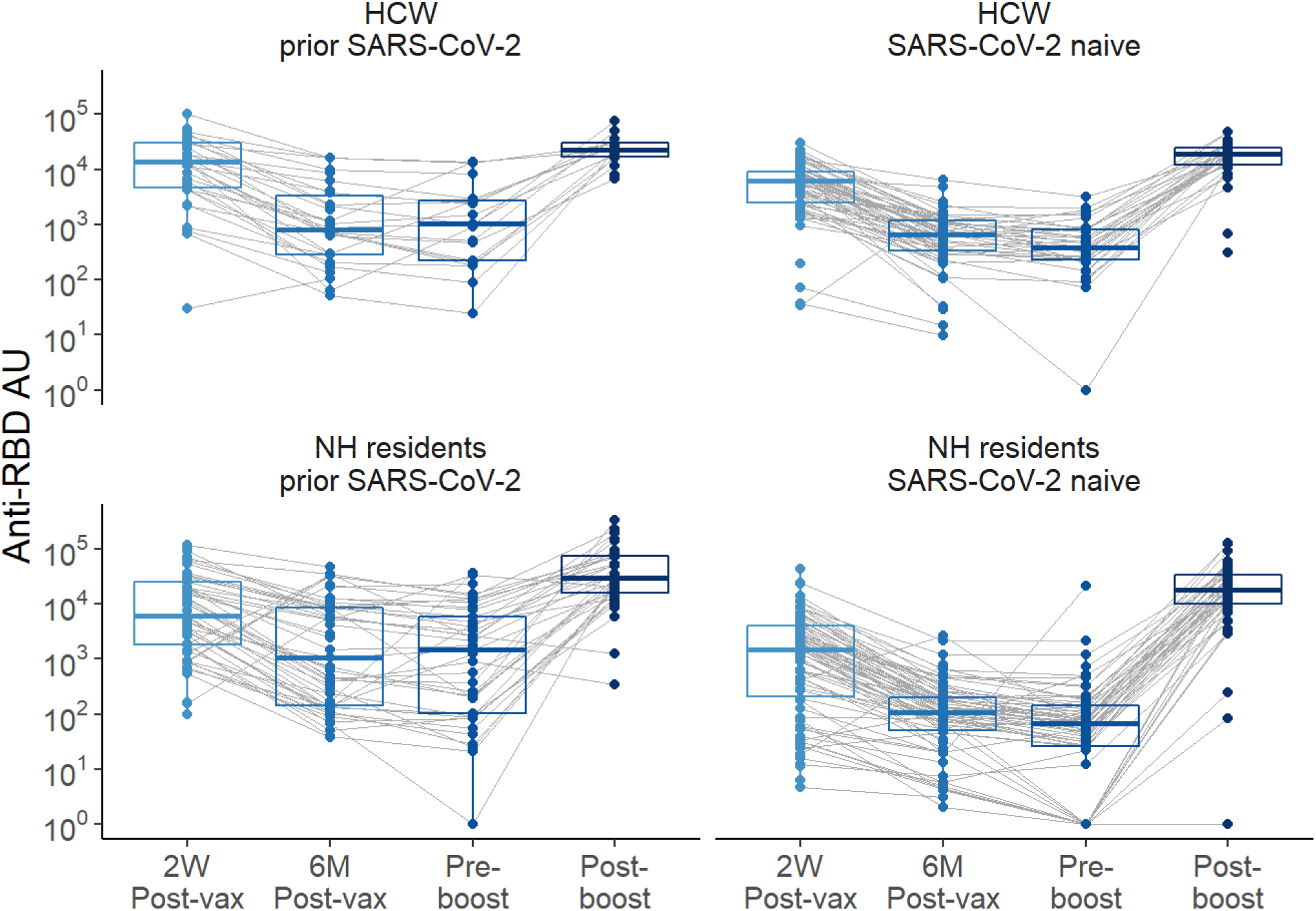
Anti-Receptor binding domain (RBD) levels over time pre- and post-boost with BNT162b2 mRNA vaccination in HCW and NH residents, with and without history of SARS-CoV-2.

Prior studies suggest increased anti-spike, anti-RBD and neutralization titers correlate with increased protection from symptomatic infection ^6,7^. Feng et al estimated that an anti-spike of 264 BAU/ml achieved 80% protection from symptomatic infection ^7^. Using the same WHO standard, we found that 95% of the NH residents reached this anti-spike level after boosting, compared with only 81% 2 weeks after the initial two-dose series. As seen in Table 2, after boosting, the magnitude above this level was substantial with anti-spike GMT increasing to 1977 BAU/ml vs 212 BAU/ml after the primary vaccine series in the infection-naive NH residents. Neutralization titers had a similar GMT increase achieving much higher levels after boosting (578 vs 96 pNT50). These significantly higher levels should allow multiple more months before they drift below the 264 BAU/ml “protective” threshold.

A fairly large proportion of infection-naive NH residents proved to be hyporesponders with low anti-spike, anti-RBD and neutralization levels after the initial 2-dose vaccine series. But the booster dose improved the hyporesponsive group’s antibody levels much closer to the median level of the responding population. These data suggest the presentation of a new neoantigen vaccine to populations who are immunologically like those living in nursing homes might benefit from much earlier receipt of a third “consolidating” dose, more akin to the three-dose strategy recommended by CDC in the immunosuppressed group. We take encouragement from the finding that most of even this debilitated and multi-morbid NH group can immunologically eventually mount a substantial antibody response to these vaccines.

Limitations of this study include the small sample size and the preponderance of males we recruited from a home for Veterans despite our efforts to resample all subjects in our prior cohort. We were unable to re-enroll subjects for a number of reasons, including refusal to obtain a booster vaccine, no longer working (HCWs) or living in the facility, and loss-to-follow-up due to non-COVID-19 interim mortality. We did not assess T-cell contribution to vaccine-induced immunity.

Given the decline in antibody levels in the 6 months following initial vaccination^1^, and the substantial increase in anti-spike, RBD and neutralization levels after booster, our data strongly support current CDC recommendations for boosting NH residents and HCWs for the likelihood that it translates to increased clinical protection. Furthermore, there are now three reports in the general population of increased clinical protection after the booster shot providing further clinical support ^8-10^.

## Data Availability

All data produced in the present work are contained in the manuscript.

## Acknowledgement

This work was supported by NIH AI129709-03S1, CDC 200-2016-91773, U01 CA260539-01, and VA BX005507-01.

## Potential conflicts of interest

S. G. and D. H. C. are recipients of investigator-initiated grants to their universities from Pfizer to study pneumococcal vaccines and Sanofi Pasteur and Seqirus to study influenza vaccines, and S.G. from Genentech on influenza antivirals. S. G. also does consulting for Seqirus, Sanofi, Merck, Vaxart, Novavax, Moderna and Janssen; has served on the speaker’s bureaus for Seqirus and Sanofi; and reports personal fees from Pfizer and data and safety monitoring board (DSMB) fees from Longevoron and SciClone. D. H. C. has done consulting work for Seqirus.

## Notes

### Funding Statement

This work was funded by NIH AI129709-03S1, CDC 200-2016-91773, U01 CA260539-01, and VA BX005507-01.

### Author Declarations

The study approval was obtained from the New England Institutional Review Board for this work.

